# Alterations in Respiratory Heart Rate Variability in Brain-Injured Neuro-ICU Patients Compared with Healthy Humans

**DOI:** 10.1101/2025.11.05.25339554

**Authors:** Valentin Ghibaudo, Gwendan Percevault, Samuel Garcia, Hugo Ardaillon, Nathalie Buonviso, Clément Menuet, Baptiste Balança

## Abstract

**Background:** Respiratory heart rate variability (RespHRV), the physiological variation in heart rate in phase with breathing, is mainly generated by central brainstem mechanisms. Its characteristics and determinants in brain-injured patients in the neuro-intensive care unit (neuro-ICU) are poorly understood.

**Objective:** To characterize RespHRV amplitude and phase in brain-injured patients compared to healthy participants, and to explore clinical variables influencing RespHRV in the neuro-ICU.

**Methods:** We analyzed 55 brain-injured patients (traumatic brain injury, aneurysmal subarachnoid hemorrhage, or other causes) and 31 healthy controls. ECG and respiratory signals were recorded and processed to extract cycle-by-cycle RespHRV amplitude and phase. Group differences were assessed using Mann–Whitney and Watson–Williams tests. In an additional analysis, 55 patients’ RespHRV amplitude and phase were modeled using generalized linear mixed-effects models to evaluate the impact of sedation, mechanical ventilation mode, vasoactive and analgesic drugs, and time, including random intercepts and slopes for subjects.

**Results:** Compared to controls, brain-injured patients exhibited a significantly lower RespHRV amplitude (1.04 [0.45, 1.96] vs. 6.21 [4.08, 9.34] bpm; p < 0.001) and an inverted RespHRV phase, with peak heart rate occurring during expiration rather than inspiration. Mixed-effects modeling revealed that machine-triggered ventilation and high level of sedation induced a significant reduction in RespHRV amplitude.

**Conclusions:** Brain-injured patients demonstrate markedly impaired central generation of RespHRV, with peripheral contributors likely accounting for the remaining variability. Ventilation mode and pharmacological interventions strongly alter RespHRV. Restoration of normal RespHRV patterns may serve as a physiological marker of autonomic and brainstem recovery, warranting further investigation in longitudinal studies.

## 1. Introduction

Respiratory Heart Rate Variability (RespHRV), recently renamed from Respiratory Sinus Arrhythmia (Menuet et al., 2025), represents one form of cardio-respiratory coupling, as heart rate normally increases during inspiration and decreases during expiration. Its mechanisms have been extensively studied since its first description in the literature (Ludwig, 1847). These mechanisms can be classified into several contributing factors, the major one being the modulation of cardiac vagal preganglionic neuron activity in the brainstem by inputs from neurons of the respiratory central pattern generator (rCPG, e.g., pre-Bötzinger Complex and Kölliker-Fuse neurons) (Berntson et al., 1993; Menuet et al., 2025; Farmer et al., 2016; Buron, 2025). The Bainbridge reflex—i.e., the increase in heart rate during inspiration mediated by veno-atrial stretch receptor feedback and direct stretch of the sinoatrial node—constitutes a minor contributor. This is evidenced by the drastic loss (>95% of the initial amplitude) of RespHRV following pharmacological, optogenetic, degenerative, surgical, or traumatic vagal de-efferentation (De Meersman, 1993; Wheeler & Watkins, 1973; Farmer et al., 2016; Buron, 2025; Menuet et al., 2020; Conci et al., 2001; Bernardi et al., 1989), which disrupts the functional link between brainstem activity and heart rate modulation.

Severe acute brain injuries encompass a heterogeneous group of conditions, including traumatic brain injury (TBI) and aneurysmal subarachnoid hemorrhage (aSAH), both requiring admission to a neuro– intensive care unit (neuro-ICU). In this setting, the optimization of vital functions and systemic homeostasis, as well as the early detection of secondary insults, are key components of critical care management. This relies on multimodal monitoring of brain function, encompassing cortical and subcortical activity. Because brainstem activity partly drives vital rhythms such as heart rate and respiratory rate, several studies have focused on its non-invasive assessment through HRV analysis (Benghanem et al., 2024; Bodenes et al., 2022; Endoh et al., 2019; Mazzeo et al., 2011; Rapenne et al., 2001; P. Zhang et al., 2020). Some of these studies have evaluated the individual components of HRV, allowing the examination of respiratory-induced heart rate variability. Some of these studies have evaluated the individual components of HRV, allowing the examination of RespHRV. However, in the neuro-ICU context, this phenomenon has almost exclusively been explored via the power in the high-frequency (HF) band using frequency domain spectral analysis (Benghanem et al., 2024; Bodenes et al., 2022; Haji-Michael et al., 2000), which provides only a coarse representation and lacks a rigorous description of the amplitude and phase of RespHRV.

Therefore, precise characterization of RespHRV may represent a valuable approach for continuous assessment of physiological states, as it could serve as a marker of brainstem function and recovery, given its central mechanisms of generation. However, the literature lacks a detailed description of RespHRV characteristics (amplitude and phase) in brain-injured patients. To address this gap, we conducted a study comparing the amplitude and phase of RespHRV in brain-injured patients hospitalized in a neuro-ICU to those in a healthy control population.

## 2. Materials and methods

### Participants

Two participant cohorts from separate studies were included: a case group comprising brain-injured patients admitted to the neuro-ICU, and a control group of healthy participants. Both studies were conducted in accordance with the Declaration of Helsinki.

#### Case group

The case group was derived from a monocentric, prospective observational trial conducted in the neurological intensive care unit (ICU) of the Hospices Civils de Lyon, France. The MultiICU study was approved by the Comité scientifique et éthique des Hospices Civils de Lyon (Ethical Committee IRB 0013204, No. 713, 06/07/2023) and registered with the French National Commission on Informatics and Liberty (CNIL No. 25_5713, France). For this observational analysis, patients or their relatives were informed about the study, and written consent was deemed unnecessary by the local ethical committee. The study investigates multiple aspects of multimodal neurological monitoring following acute brain injuries, including aneurysmal subarachnoid hemorrhage (SAH), severe traumatic brain injury (TBI), and intraparenchymal hematoma. Data were collected from digital medical records starting in January 2020.

#### Control group

The control group was obtained from a previously published study (Ghibaudo et al., 2025). In this study, thirty-one participants attended two sessions at the Lyon Neuroscience Research Center (Registration number DPO service: 2-20125). Inclusion criteria were: age between 18 and 60 years, social security coverage, and willingness to participate. Exclusion criteria included pregnancy, labor, or breastfeeding; deprivation of liberty by judicial or administrative decision; and known cardiovascular, respiratory, olfactory, neurosensory, or psychiatric disorders. The study was approved on May 17, 2022, by the Comité de Protection des Personnes Ile-de-France 3 (ID RCB: 2021-A03077-34), in accordance with French regulations for biomedical research involving healthy volunteers. Participants were clearly and fairly informed about the study, provided written informed consent, and were compensated for their participation. During the second session, physiological, neuronal, and psychological data were recorded across three experimental conditions. The first condition corresponded to a “baseline” state, in which participants were asked to sit comfortably in a chair with their eyes open.

### Data acquisition

#### Case group

Data were collected using the Moberg CNS Monitor (CNS-320, MICROMED-NATUS, USA). Respiratory activity was measured via end-tidal CO₂ using an Infinity® MCable™ Mainstream CO₂ sensor (Dräger Médical, Antony, France). Electrocardiographic (ECG) signals were recorded with a 5-lead thoracic system. All signals were continuously recorded and displayed on the IntelliVue MX750 monitor (Philips, Suresnes, France) connected to the CNS monitor, with sampling rates of 500 Hz for ECG and 60–120 Hz for CO₂. Continuous clinical and therapeutic data were also extracted from the IntelliSpace Critical Care and Anesthesia (ICCA, Philips, Suresnes, France) software, which is routinely used in the unit for prescription and patient management.

#### Control group

Respiratory signals were recorded using nasal cannulas connected to a pressure sensor (Sensortechnics GmbH, Puchheim, Germany) that captured variations in nasal airflow. Cardiac activity was recorded via an electrocardiogram (ECG, Brain Products GmbH, Gilching, Germany) with three electrodes placed on the anterior surface of the right wrist, the left wrist, and the lower abdomen at the left iliac fossa. Both respiratory and ECG signals were acquired using a DC amplifier (actiCHamp Plus, Brain Products GmbH, Gilching, Germany) at a sampling rate of 1000 Hz.

### Data processing

Data processing was performed using Python (version 3.12). All code used in the analyses is available via the link provided in the Code Availability statement at the end of the manuscript.

#### Case group: from raw files to python using *pycns* toolbox

For the case group, we developed a custom Python toolbox, *pycns* (available at https://github.com/samuelgarcia/pycns), specifically designed to convert raw Moberg CNS Monitor files into a Python-compatible format. This toolbox enabled the generation of continuous CO₂ and ECG time series for each patient in NumPy format (Harris et al., 2020), facilitating efficient scientific computation and analysis.

#### Control group: from raw files to python using *physio* toolbox

For the control group, raw data were converted to Python NumPy format using the *physio.read_one_channel()* function from the *physio* toolbox (Ghibaudo et al., 2023).

#### Computing amplitude of RespHRV using *physio* toolbox

The processing of ECG and respiration data to obtain a precise, cycle-by-cycle measure of RespHRV using the *physio* toolbox has been described in a dedicated publication (Ghibaudo et al., 2023), where all details are provided. Additional information is available in the toolbox documentation: https://physio.readthedocs.io/en/latest/examples/example_05_resphrv.html. For clarity, we summarize here only the main steps.

##### 1. Respiration

Detection of respiratory cycles (i.e., expiration-inspiration and inspiration-expiration transitions) was performed using the *physio.compute_respiration()* function. For the case group, the raw CO₂ signal was processed with the preset *human_co2*, and for the control group, the raw airflow signal was processed with the preset *human_airflow*. These parameter presets consist of optimized settings controlling several processing steps, including filtering, cycle detection, and removal of aberrant cycles.

##### 2. ECG

Detection of ECG R peaks was performed using the *physio.compute_ecg()* function, with the raw ECG signal processed using the preset *human_ecg* for both groups. This preset includes optimized parameters for filtering, peak detection, and cleaning of aberrant peaks.

##### 3. RespHRV amplitude

Heart rate dynamics were computed for each respiratory cycle by providing the detected respiratory cycles and ECG R peaks to the *physio.compute_resphrv()* function. Among the various features obtained, we retained only the difference between the maximum and minimum heart rate within each respiratory cycle, corresponding to the amplitude of the respiratory-induced heart rate variability.

#### Computing phase of RespHRV using *physio* toolbox

In healthy subjects, an increase in heart rate during inspiration and a decrease during expiration is expected. However, an inversion of this typical relationship has been reported in fMRI studies (Rassler et al., 2022), which the authors attributed to altered serotonergic and noradrenergic activity associated with elevated anxiety. Thus, the direction of phasing may depend on brain physiology, suggesting that this parameter could serve as a potential marker of brain recovery in brain-injured patients. We therefore developed an analysis to compare this relationship between our case and control groups. First, an instantaneous heart rate vector was computed from RR intervals. Second, detected respiratory cycles were used to cyclically deform the instantaneous heart rate trace, rescaling its time vector into a respiratory phase basis to construct a 2D *resp cycle × resp phase* matrix of heart rate. Finally, this matrix was averaged across respiratory cycles to obtain the mean heart rate as a function of respiratory phase.

##### 1. Instantaneous heart rate

Previously detected ECG R peaks were provided to *physio.compute_instantaneous_rate()*, which re-interpolated the RR intervals to a regularly sampled time series matching the original ECG trace using linear interpolation. The resulting values were then converted into beats per minute (60 / RR_i_ [s]), generating an instantaneous heart rate vector sampled at the same rate as the raw ECG signal.

##### 2. Cyclical deformation of heart rate trace

The function *physio.deform_to_cycle_template()* was used to rescale the time basis of the instantaneous heart rate vector according to the respiratory cycle timepoints previously detected, using linear interpolation. In this way, a 2D matrix was obtained for each subject, with dimensions (*n_respiratory_cycles × n_respiratory_phase_points*), where each value represents the heart rate (see documentation of the *physio* toolbox for a graphical view of such matrix: https://physio.readthedocs.io/en/latest/examples/example_05_resphrv.html). The number of phase bins per respiratory cycle was set to 100 for both groups, with bins 1–40 corresponding to inspiration and bins 41–100 corresponding to expiration.

##### 3. Averaged heart rate by respiratory phase bin

The 2D matrix of shape (*n_respiratory_cycles × n_respiratory_phase_points*) was then averaged across cycles using the median, yielding a single trace per subject of shape (*n_respiratory_phase_points*) that describes the average heart rate as a function of respiratory phase. This resulted in a matrix of shape (*n_subjects × n_respiratory_phase_points*) for each group.

### Data segmentation

The recordings from the control group lasted 10 minutes. To achieve equivalent statistical power, we selected a 10-minute segment from each patient in the case group, extracted from several days of continuous monitoring. This segment corresponded to the first ten minutes of the first hour after one full day of recording for each patient, which occurred on a median of 2.81 [1.93, 3.71] days after hospital admission. We chose this recording time to potentially maximize variability in the data, as some patients had partially recovered brain function, and to balance the number of patients triggering their own respiration with those fully dependent on the ventilator (Figure 2A and C).

### Additional analysis in the case (patient) group

Since this study aimed to investigate RespHRV in brain-injured patients, we conducted an additional analysis to identify variables that could influence RespHRV in the neuro-ICU context. Indeed, factors such as sedation level, vasoactive medications, ventilation mode (assisted or fully controlled), analgesic drugs, and time could potentially modulate RespHRV. To examine the co-evolution of these variables with RespHRV, we reconstructed regularly spaced time series for each variable, aligned to the patient’s hospital stay, with a 10-minute sampling interval, using the following method.

#### RespHRV amplitude

As described previously, the amplitude of RespHRV was computed for each respiratory cycle. The reconstructed time series corresponded to the median RespHRV amplitude of all respiratory cycles detected within each 10-minute time interval, spanning from the beginning to the end of the recording.

#### RespHRV phase

The respiratory phase corresponding to each patient’s maximum heart rate (HR) was identified by determining whether the maximal HR occurred during inspiration or expiration, based on the averaged heart rate profile over the respiratory cycle within each 10-minute time interval, spanning the entire recording. Boundary phases were excluded to avoid ambiguity (2.5% margin at the edges of the respiratory phase). This procedure yielded a 10-minute time series of labels, “inspiration” or “expiration,” indicating whether the average maximum HR occurred during inspiration or expiration, respectively.

#### Ventilation mode

Ventilation mode in our data can be classified in two ways: fully controlled, where the ventilator initiates inspiration and provides mechanical inflation of air into the patient’s lungs, and assisted, where the patient initiates the inspiration effort and the ventilator provides mechanical support. During fully controlled ventilation, respiratory variability is nearly absent, whereas during assisted ventilation, natural variability is preserved and generally higher. Therefore, ventilation mode was inferred from the local variability of respiratory cycle durations detected within each 10-minute time interval. Periods were classified as “machine-triggered” (i.e. fully controlled) when variability was near zero and as “patient-triggered” (i.e. assisted) when variability was higher. An automated threshold was computed for each subject to distinguish the two regimes, defined as the first minimum in the histogram of the rolling standard deviation of respiratory cycle durations.

#### ICCA software data (sedation, vasoactive, analgesic)

All therapeutic doses recorded in the IntelliSpace Critical Care and Anesthesia software (ICCA, Philipps©, Netherlands) were collected hourly and during bolus from the automatic pump and entered by caregivers at irregular time intervals for other medications. These data were resampled at regular 10-minute intervals using interpolation with *scipy.signal.interp1d* (Virtanen et al., 2020) and the *previous* method. This approach was preferred over linear interpolation, which could have inferred inaccurate doses, given that the pharmacodynamics of continuous infusion are intended to maintain as constant a delivery as possible.

For sedation level, a composite score was constructed by counting the number of sedative drugs administered at each time point, among propofol, midazolam, and thiopental. The score ranged from 0 for no sedatives, 1 for a single drug, and 2 for two or more drugs. Because thiopental alone has a profound sedative effect, its administration alone was assigned a score of 2.

For vasoactive and analgesic drugs, only the doses of noradrenaline and sufentanil were retained, expressed in µg/h/kg and mg/h/kg, respectively, and sampled at 10-minute intervals

The resulting time series for each patient were cropped to maximize the period of simultaneous availability of all variables. For this reason, remifentanil was not included, as too few patients received this drug—its inclusion would have reduced the analysis to fewer than 36 patients.

### Statistics

#### Group analysis

RespHRV amplitude was compared between the case group (brain-injured patients) and the control group (healthy participants) using a two-sided, non-parametric Mann–Whitney U test, implemented via the *pingouin.mwu* function from the Pingouin toolbox. The null hypothesis (H₀) was rejected when the p-value was less than 0.05. A subgroup analysis was also performed within the patient group to compare RespHRV amplitude between those able to trigger their own respiration (*patient-triggered* respiration) and those fully dependent on mechanical ventilation (*machine-triggered* respiration), using the same statistical procedure (results shown in Figure 2A).

For RespHRV phase, the 2D matrix of shape (*n_subjects × n_respiratory_phase_points*), where *n_subjects* represents the number of participants in each group, was reduced to a 1D vector of shape (*n_subjects*) by computing the mean angular direction of each subject’s phase histogram using *pingouin.circ_mean()* from the *pingouin* toolbox. This yielded one vector of mean RespHRV phasing angles per group. These vectors were then analyzed using the Watson–Williams test via *pycircstat.tests.watson_williams()* from the *pycircstat* toolbox to determine whether the average phase direction differed significantly between groups. Differences were considered statistically significant if the p-value was below 0.05. Furthermore, the respiratory phase of the maximum heart rate (HR) corresponding to each participant was identified by determining whether the maximal HR occurred during inspiration or expiration, based on the averaged heart rate profile over the respiratory cycle. Boundary phases were excluded to avoid ambiguity (2.5% margin of exclusion at the edges of the respiratory phase). The distribution of these labeled phases (maximum HR during *inspiration* vs. *expiration*) was then compared between the control and patient groups using a chi-square (χ²) test of independence (*pingouin.chi2_independence* function, Pingouin toolbox) to assess whether the likelihood of exhibiting a maximum HR during inspiration or expiration differed between groups (results shown in Figure 2B). The same statistical procedure was applied to the patient subgroup to compare the likelihood of exhibiting a maximum HR during inspiration or expiration according to ventilatory mode (*patient-triggered* vs. *machine-triggered* ventilation) (results shown in Figure 2C).

#### Additional analysis in the case (patient) group

As described previously, an additional analysis was conducted to explore the co-evolution of clinical variables on RespHRV amplitude in brain-injured patients in the neuro-ICU. This analysis employed a multivariate regression model aimed at predicting RespHRV amplitude using fixed-effect variables: sedation level (categorical: 0, 1, 2), ventilation mode (categorical: controlled, assisted), noradrenaline dose (continuous), sufentanil dose (continuous), and time (continuous). Given the high inter-individual variability of RespHRV amplitude, patient-specific intercepts were included as a random effect. Additionally, random slopes for sufentanil and time were incorporated, allowing the effects of these variables to vary across individuals. This structure was selected after comparing alternative specifications: including additional random slopes, such as for noradrenaline, resulted in negligible variance estimates and did not improve model fit, indicating that inter-individual variability for these covariates was minimal.

Since RespHRV amplitude data are Gamma distributed, the model was specified to account for Gamma-distributed residuals. The model was implemented using the *glmmTMB()* function from the *glmmTMB* package in R (version 4.4.2), corresponding to a generalized linear mixed-effects model. The model formula was specified as follows:

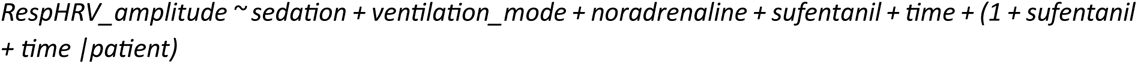

Model estimates were extracted, including β-coefficients, 95% confidence intervals (CIs), and p-values. Significance levels were categorized as p ≤ 0.001 (***), p ≤ 0.01 (**), p ≤ 0.05 (*), and non-significant (ns). To aid interpretation, fixed-effect estimates from the Gamma log-link model were exponentiated and expressed as percent changes in RespHRV amplitude using the formula (exp(β) – 1) × 100, where β represents the model coefficient on the log scale (Table 1).

**Table 1.**
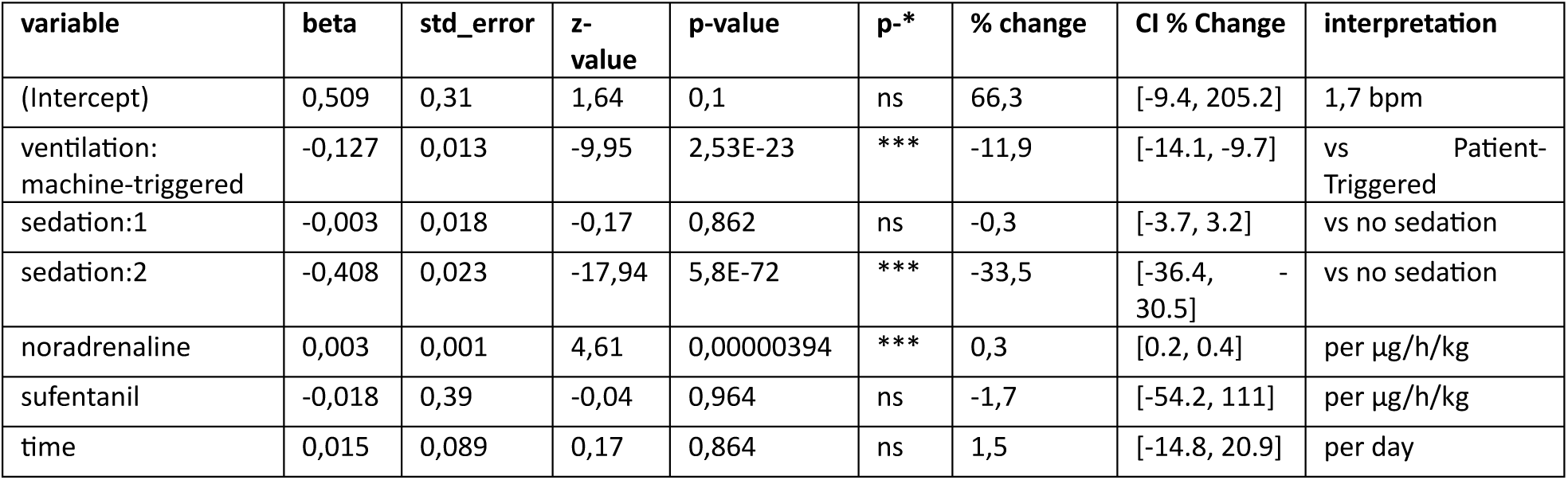
Results of generalized mixed linear model explaining RespHRV amplitude by clinical variables.

Furthermore, we performed an equivalent statistical analysis to explore the effects of clinical variables on the RespHRV phase in these patients. As described above, we constructed a 10-minute time series of labels, “inspiration” or “expiration,” indicating whether the average maximum HR occurred during inspiration or expiration, respectively, spanning the entire recording. This allowed us to analyze the probability of the maximum HR occurring during inspiration or expiration according to clinical variables, using a binomial generalized linear mixed model (GLMM) (*glmmTMB* function / package, binomial family, logit link) specified as follows:

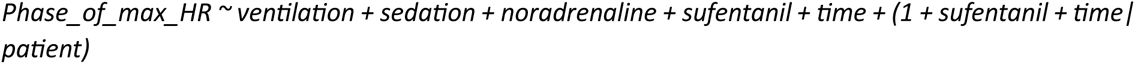

Model estimates were extracted, including β-coefficients, 95% confidence intervals (CIs), and p-values. Significance levels were categorized as p ≤ 0.001 (***), p ≤ 0.01 (**), p ≤ 0.05 (*), and non-significant (ns). To aid for interpretation, fixed-effects estimates from the binomial beta estimators obtained with a logit link were exponentiated to obtain odds ratios (ORs), using the formula OR = exp (*β*), where *β* represents the model coefficient on the logit scale. An OR greater than 1 indicates an increase in the odds of the event of interest (e.g., the maximum of heart rate occurring during inspiration) per unit increase in the predictor, whereas an OR less than 1 indicates a decrease in the odds. For categorical predictors, the OR represents the odds relative to the reference category. For continuous predictors, the OR represents the multiplicative change in odds per unit increase. Probabilities of the event can be derived from the OR using 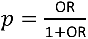 for the reference condition.

## 4. Results

### Dataset description

#### Case group

The main study included 55 brain-injured patients from 2020 to 2025 (31 males, 23 females, 1 missing data), with a median age of 48 years [39, 57] (median [1st quartile, 3rd quartile]). This number of 55 patients results from an initial cohort of 98 eligible patients, of whom 33 were excluded due to missing or poor-quality etCO₂ and ECG data during the period of analysis. Patients were admitted to the neuro-ICU for aneurysmal subarachnoid hemorrhage (SAH, N = 28), traumatic brain injury (TBI, N = 20), or other causes (N = 7). The initial Glasgow Coma Scale (GCS) score was 3 [3, 3], and the final GCS score was 11 [9, 14].

#### Control group

The main study included 31 healthy participants (16 females, 15 males), with a median age of 26 years [22, 32].

### Main results

RespHRV amplitude was significantly lower in the case group (1.43 [0.7, 2.99] bpm) compared to controls (6.27 [4.09, 8.64] bpm; U = 150, p < 0.001; Figure 1A). Median heart rate did not differ significantly between controls (74.07 [68.15, 81.17] bpm) and cases (77.68 [65.83, 87.81] bpm; U = 944, p = ns). Heart rate variability (HRV, computed as median absolute deviation) was significantly higher in controls (5.34 [3.89, 6.51] bpm) than in cases (1.64 [1.02, 3.56] bpm; U = 229, p < 0.001). Respiratory rate was significantly lower in controls (14.05 [11.32, 16.32] cpm) than in cases (18.28 [16.0, 20.04] cpm; U = 1390, p < 0.001), while respiratory rate variability (RRV, median absolute deviation) was significantly higher in controls (1.79 [1.4, 2.29] cpm) than in cases (0.38 [0.17, 0.65] cpm; U = 194, p < 0.001). During the analysis period, the median Glasgow Coma Scale score was 15 [15, 15] in controls and 3 [3, 3] in the case group (U = 0, p < 0.001). A subgroup analysis comparing RespHRV amplitude between patients under mechanical ventilation triggered by their endogenous respiratory activity (*patient-triggered* respiration) and those with mechanical ventilation triggered by the ventilation machine (*machine-triggered* respiration) showed a tendency for a difference that did not reach statistical significance (U = 230, p = 0.07) (Figure 2A). Given the effect of respiratory frequency on RespHRV amplitude, we examined whether it differed between the two subgroups; no significant difference was found: machine-triggered 18.0 [15.74–20.01] cpm vs. patient-triggered 19.12 [16.14–21.28] (p = 0.16).

**Figure 1.**
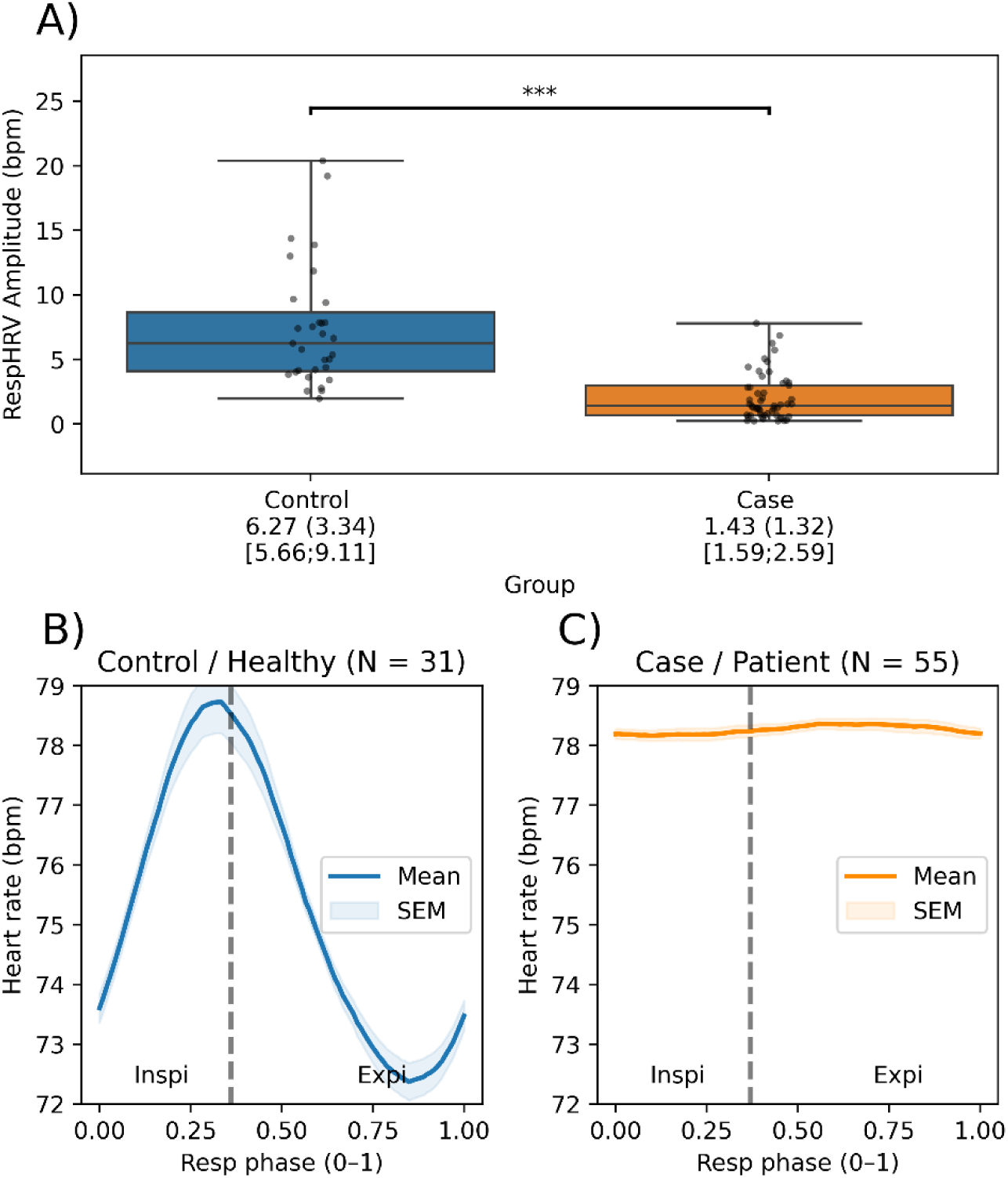
A) Comparison of RespHRV amplitude (in beats per minute, bpm) between Control (healthy participants, N = 31) and Case (brain-injured patients, N = 55) groups. Values for each group are presented as: Median (MAD) [95% CI]. and C) RespHRV phase between Control and Case groups, respectively.

**Figure 2.**
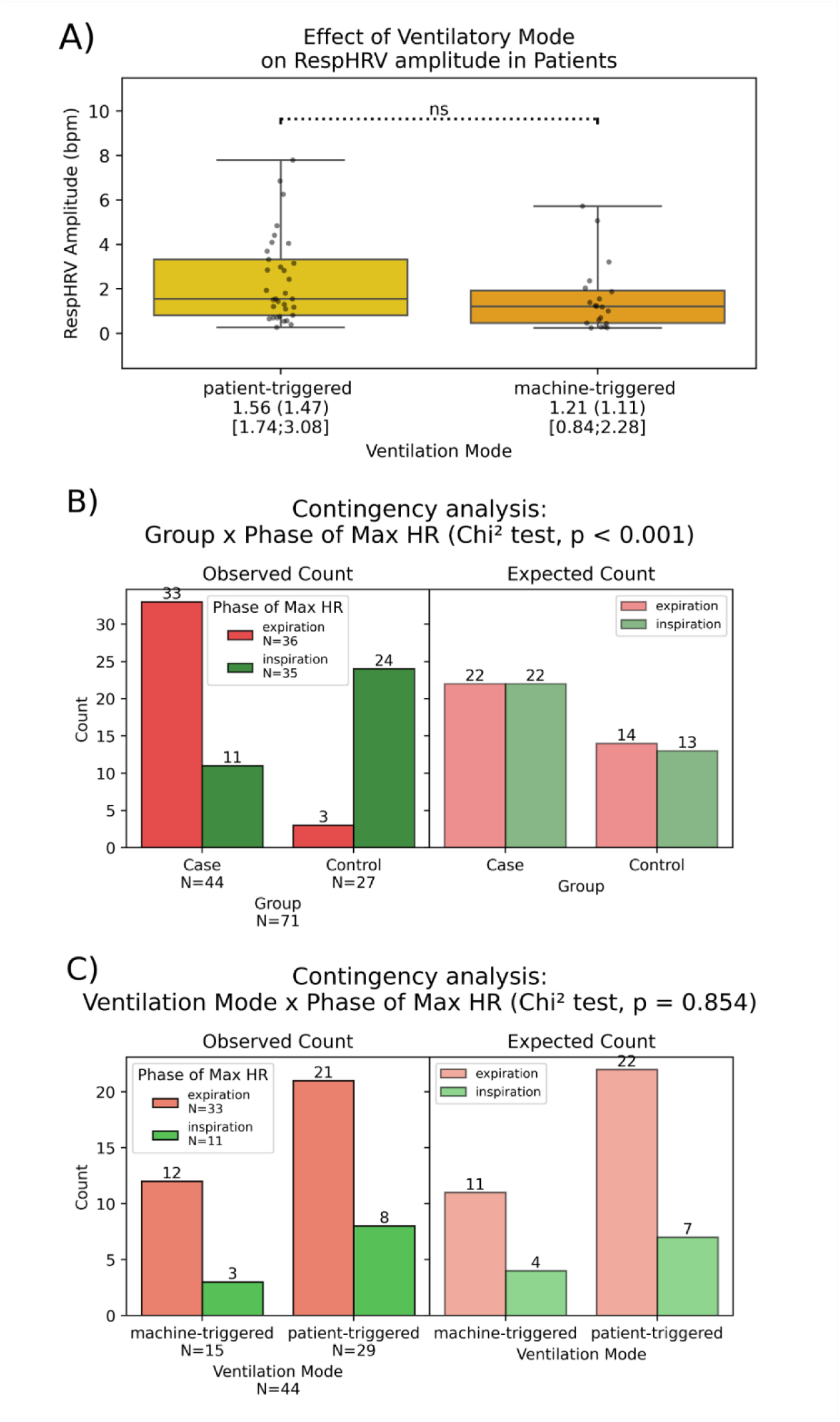
A) RespHRV amplitude (beats per minute, bpm) in patients with mechanical ventilation that is triggered by their endogenous respiratory activity (patient-triggered, N = 35) or by the ventilation machine (machine-triggered, N = 20. Note that “ns” stands for non-significant; however, the result shows a trend that did not reach statistical significance (U = 230, p = 0.07). B) RespHRV phase: Contingency analysis of subject group (Control or Case) versus the predominant phase of maximum heart rate (Phase of Max HR). The left panel shows the observed counts of subjects per Group/Phase, and the right panel shows the expected counts under the assumption of no association between the variables. C) RespHRV phase: Contingency analysis of ventilatory mode (patient- or machine-triggered) versus the predominant phase of maximum heart rate (Phase of Max HR).

Moreover, RespHRV phasing differed significantly between groups (Watson–Williams test: F = 109, p < 0.001), with the peak heart rate occurring during inspiration in controls and during expiration in cases (Figures 1B and 1C, respectively). After binarizing the phases to assess whether the likelihood of exhibiting a maximum HR during inspiration or expiration differed between groups, a chi-square test of independence revealed a statistically significant effect of group on the phase of maximum heart rate (p < 0.001; Figure 2B). Applying the same test to the patient subgroup, no significant effect of ventilatory mode (patient-triggered vs. machine-triggered) on the phase of maximum heart rate was observed (p = 0.85; Figure 2C).

### Additional analysis

This additional analysis included 55 patients (37 males, 17 females, 1 missing data) with a median age of 47 years [36, 56]. Patients were admitted to the neuro-ICU for traumatic brain injury (TBI, N = 25), aneurysmal subarachnoid hemorrhage (SAH, N = 23), or other causes (N = 7).

RespHRV amplitude was analyzed using a generalized linear mixed-effects model with a Gamma distribution and log link (see Table 1). Among categorical covariates, sedation level 2 was associated with a 33.5 % decrease in RespHRV compared to no sedation (β = -0.408, p < 0.001; 95 % CI for % change: [-36.6 %, -23.0 %]), whereas sedation level 1 had no significant effect (β = -0.003, p = 0.862; % change ≈ -0.3 %, 95 % CI: [-3.5 %, 3.1 %]). Machine-triggered ventilation showed a small but highly significant decrease in RespHRV compared to patient-triggered ventilation (β = -0.127, p < 0.001; % change ≈ -11.9 %, 95 % CI: [-13.9 %, -9.7 %]). For continuous covariates, noradrenaline was associated with a modest but statistically significant positive effect (β = 0.003, p < 0.001; % change ≈ +0.3 %, 95 % CI: [0.2 %, 0.4 %]), while sufentanil and time showed no significant effects (p = 0.964 and p = 0.864, respectively). The random effect of subject captured substantial inter-individual variability (ICC = 0.950). Overall, the model explained 95 % of the variance when including both fixed and random effects (conditional R²), whereas fixed effects alone accounted for 0.5 % of the variance (marginal R²). The residual dispersion of the Gamma model (σ) was 0.718. These results indicate that RespHRV amplitude is strongly influenced by sedation level 2 and ventilatory mode, with smaller contributions from noradrenaline, while sufentanil and time have no effects.

A generalized linear mixed-effects model with binomial distribution and logit link was used to assess the likelihood of the maximum heart rate (HR) occurring during inspiration (see Table 2). Machine-triggered ventilation significantly increased the probability of maximum HR during inspiration (β = 0.426, SE = 0.046, z = 9.17, p < 0.001; OR = 1.53 [95 % CI: 1.40, 1.66]). Sedation level 1 decreased the likelihood (β = -0.639, SE = 0.067, z = -9.49, p < 0.001; OR = 0.53 [95 % CI: 0.46, 0.61]), whereas sedation level 2 had a smaller but still significant effect (β = -0.487, SE = 0.084, z = -5.81, p < 0.001; OR = 0.61 [95 % CI: 0.52, 0.71]). Among continuous covariates, noradrenaline had a modest but significant negative effect (β = -0.006, SE = 0.002, z = -2.32, p = 0.021; OR = 0.994 per µg/h/kg [95 % CI: 0.990, 0.998]), while sufentanil and time had no significant effects. The random effect of subject captured substantial inter-individual variability (ICC = 0.951), and the conditional R² of the model was 0.951. These results indicate that ventilatory mode is the primary determinant of the respiratory phase in which maximum HR occurs, with machine-triggered ventilation favoring inspiration. Sedation and noradrenaline exert secondary modulatory effects, whereas sufentanil and temporal factors have minimal influence.

**Table 2.** Results of the generalized linear mixed-effects model explaining RespHRV phase (the probability of a heart rate peak occurring during inspiration) in relation to clinical variables. OR = odds ratio. An OR > 1 indicates an increased probability of a heart rate peak occurring during inspiration, an OR < 1 indicates a decreased probability (i.e., an increased probability of a peak occurring during expiration), and an OR = 1 corresponds to a 50% probability.

| variable | beta | std_error | z-value | p-value | p-* | OR | CI OR | interpretation |
| --- | --- | --- | --- | --- | --- | --- | --- | --- |
| (Intercept) | 1,368 | 1,21 | 1,13 | 0,258 | ns | 3,927 | [0.367, 42.076] | OR > 1 = inspiration |
| ventilation:<br>machine-triggered | 0,426 | 0,046 | 9,17 | 4,62E-20 | *** | 1,53 | [1.397, 1.676] | vs Patient-Triggered |
| sedation:1 | -0,639 | 0,067 | -9,49 | 2,29E-21 | *** | 0,528 | [0.463, 0.602] | vs no sedation |
| sedation:2 | -0,487 | 0,084 | -5,81 | 6,14E-09 | *** | 0,614 | [0.521, 0.724] | vs no sedation |
| noradrenaline | -0,006 | 0,002 | -2,32 | 0,0205 | * | 0,994 | [0.99, 0.999] | per µg/h/kg |
| sufentanil | 0,644 | 2,439 | 0,26 | 0,792 | ns | 1,904 | [0.016, 226.963] | per µg/h/kg |
| time | 0,007 | 0,283 | 0,02 | 0,982 | ns | 1,007 | [0.578, 1.751] | per day |

Together, these analyses indicate a dissociation between factors affecting the magnitude versus the phasing of respiratory-related heart rate fluctuations. Sedation level 2 and ventilatory mode substantially modulate RespHRV amplitude, while ventilatory mode is the strongest determinant of the timing of maximum HR, with machine-triggered ventilation shifting it toward inspiration.

## 5. Discussion

We investigated RespHRV in brain-injured patients hospitalized in the neuro-ICU compared to healthy participants and observed two main alterations: (1) a markedly reduced RespHRV amplitude and (2) a shift in RespHRV phasing in patients. Further analysis using generalized linear mixed-effects models indicated that, after adjusting for other variables, high sedation levels were the primary factor reducing RespHRV amplitude, whereas ventilation mode was the main determinant of RespHRV phasing.

These findings suggest that central mechanisms generating RespHRV are strongly impaired in brain-injured patients under neuro-ICU care, leaving only minor peripheral contributors that normally play a small role in physiology. This aligns with the loss of RespHRV amplitude observed following pharmacological (e.g., muscarinic antagonists), optogenetic (e.g., modulation of respiratory inputs to cardiac vagal neurons), degenerative (e.g., diabetic dysautonomia), surgical (e.g., cardiac transplantation), or traumatic (e.g., brainstem death) vagal de-efferentation, which disrupts the functional link between brainstem activity and heart rate modulation (Bernardi et al., 1989; De Meersman, 1993; Wheeler & Watkins, 1973; Farmer et al., 2016; Menuet et al., 2020; Buron, 2025; Conci et al., 2001). Worth noting, in the patient group, after adjustment for all clinical variables, the intercept corresponded to a RespHRV amplitude of 1.8 bpm—much lower than that observed in controls—suggesting a substantial effect of the brain injury itself on the reduction of RespHRV. Several mechanisms may contribute to this phenomenon, although all remain speculative and require dedicated investigation in experimental animal models of traumatic brain injury. First, a marked rise in intracranial pressure may reduce cerebral perfusion pressure and precipitate transtentorial herniation, both resulting in brainstem compression and subsequent disruption of critical functions, including respiratory rhythmogenesis (Benghanem et al., 2020; Bragin et al., 2013; Canac et al., 2020; Y. Zhang et al., 2024). Second, injury affecting components of the central autonomic network (notably the insular cortices) could impair descending telencephalic projections to brainstem autonomic nuclei, thereby altering the interactions between respiratory and cardiovascular autonomic control (Sykora et al., 2016; Takahashi et al., 2015; Williamson et al., 2013). Third, neuroinflammation triggered by the primary lesion may induce the release of damage-associated molecular patterns, amplify oxidative stress, increase the likelihood of initiating spreading depolarizations, and ultimately compromise brainstem function if sufficiently intense to affect these structures (Balança et al., 2021; Richter et al., 2010). Besides, while both pharmacologic and traumatic factors could explain the drastic loss of RespHRV amplitude in the brain-injured patients in the present study, the reason for the inverted RespHRV phasing remains to be clarified. In normal physiology, RespHRV phasing is characterized by an increased heart rate during inspiration, largely due to inhibitory GABAergic and glycinergic inputs from the pre-Bötzinger Complex to the cardiac vagal preganglionic neurons of the nucleus ambiguus, which discharge during inhalation and decrease parasympathetic tone, thereby raising heart rate (Buron, 2025; Menuet et al., 2020). Considering patient physiology and these mechanisms, one might expect that a change from fully controlled ventilation (machine-triggered inspiration) to assisted ventilation (patient-triggered inspiration) would restore normal RespHRV phasing, assuming recovery of normal respiratory central pattern generator function. However, in our study, periods when patients underwent such a change in ventilatory mode showed only a slight increase in RespHRV amplitude (cf. Table 1), while RespHRV phasing remained for most of the patients inverted. This suggests persistent low power or desynchronization (Potts & Paton, 2006) of central brainstem mechanisms generating RespHRV compared to peripheral contributors such as the Bainbridge reflex, which may invert in patients under positive-pressure ventilation, where inspiration increases intrathoracic pressure rather than decreasing it as in spontaneous breathing (Crystal & Salem, 2012). Our results are consistent with fMRI studies reporting inverted RespHRV phasing attributed to altered serotonergic and noradrenergic activity associated with elevated anxiety (Rassler et al., 2022), which may also be altered in neuro-ICU patients.

The loss of central RespHRV generation in neuro-ICU patients may be further reinforced by therapeutic agents. Our multivariate analysis showed a strong impact of high sedation levels (tier 2, i.e., two or more sedative drugs among thiopental, midazolam, propofol, or thiopental alone), resulting in an almost 34% loss of RespHRV amplitude possibly due to pharmacological disconnection between brainstem vagal centers and the sinoatrial node. Sedative agents also decreased the probability of normal RespHRV phasing, producing an inverted pattern (maximum HR during expiration), effectively replicating the inverted RespHRV pattern previously observed in urethane-anesthetized rats (Bouairi et al., 2004). Considering these factors, we created a hypothetical figure illustrating the potential suppression of central mechanisms generating RespHRV in neuro-ICU patients compared to healthy participants (see Figure 3). However, given the exploratory nature of our study, future investigations using carefully controlled invasive experiments in animal models are warranted to disentangle the specific effects and underlying mechanisms of sedatives, noradrenaline, opioids, and different modes of artificial ventilation on both central autonomic and peripheral generation of RespHRV, as well as on the activity of the neurons of the respiratory central pattern generator.

**Figure 3.**
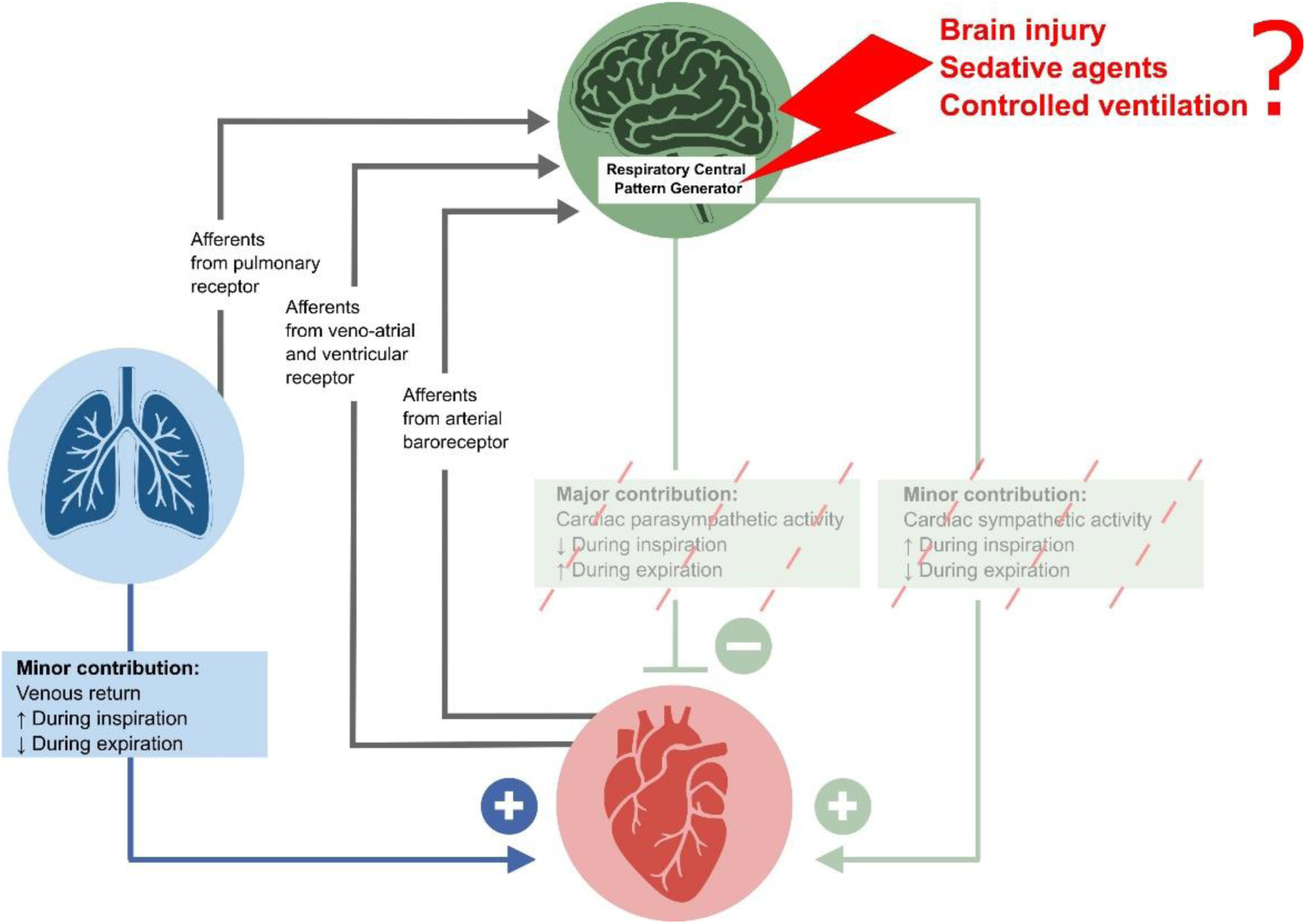
Hypothesis: loss of major central contributors in the generation of respiratory heart rate variability (RespHRV) in brain-injured patients. These alterations may explain the low amplitude and negative phasing of the RespHRV observed, the latter likely being driven by positive inspiratory pressure during artificial ventilation. Figure inspired by Menuet et al., 2025.

However, the present study has several limitations. First, the RespHRV amplitude measured was not normalized by respiratory frequency or tidal volume—two respiratory parameters known to strongly influence RespHRV (Hirsch & Bishop, 1981). Nonetheless, given that the respiratory pattern in the Neuro-ICU patients was partially or fully controlled by the ventilator with tidal volume between 6 and 8 ml/kg, these parameters likely showed minimal variation and therefore had a limited impact on the results. Moreover, the effect of breathing frequency on RespHRV amplitude follows two linear regimes when expressed in log-space, with a corner frequency around 7 cycles per minute under normal physiological conditions (Hirsch & Bishop, 1981). This relationship may be altered in brain-injured patients under Neuro-ICU care and could potentially serve as a marker of autonomic function in this context. Then, we did not adjust the RespHRV amplitude analysis for age, although there was a clear age difference between groups: the case group was on average 46 years old, whereas the control group averaged 28 years old. The literature consistently shows that RespHRV amplitude decreases with age (Hellman & Stacy, 1976; Hrushesky et al., 1984; Jennings & Mack, 1984). However, this decline is described as approximately 10% per decade (Hrushesky et al., 1984). Consequently, the expected age-related reduction over an 18-year difference would be relatively small and therefore cannot account for the large effect size observed in our data (6.27 bpm vs. 1.43 in the control and case groups, respectively). This supports the interpretation that factors other than age play a predominant role in the substantial RespHRV attenuation observed in the case group. Lastly, RespHRV was not compared with other HRV frequency bands and was not used as a predictor of clinical outcome, which may influence its amplitude and phasing. Previous studies have emphasized the relative contributions of low- and high-frequency sources of HRV, including respiratory components, for predicting outcomes in neuro-ICU patients (Baillard et al., 2002; Benghanem et al., 2024; Bodenes et al., 2022; Haji-Michael et al., 2000; Mazzeo et al., 2011; Rapenne et al., 2001). These studies generally relied on global spectral analysis of HRV, lacking the precise temporal and phasing information that a respiratory cycle-by-cycle analysis of heart rate dynamics can provide.

Considering the mechanisms identified in the present study and the features explored, future studies incorporating such cycle-by-cycle analysis of heart rate dynamics may enhance the predictive value of cardiac and respiratory recordings for clinical recovery. Restoration of normal RespHRV patterns could potentially serve as a marker of autonomic or brainstem recovery, providing clinically useful guidance for therapeutic decisions, such as the timing of extubation.

## Conclusion

In this study, we investigated respiratory heart rate variability (RespHRV) in brain-injured patients hospitalized in the neuro-ICU compared to healthy participants. We observed two main alterations: markedly reduced RespHRV amplitude and an inverted RespHRV phasing. These findings indicate a profound impairment of central brainstem mechanisms generating RespHRV, with residual variability likely driven by peripheral contributors such as the Bainbridge reflex. Ventilation modes and sedative agents also strongly alter RespHRV. Restoration of normal RespHRV patterns may serve as a sensitive physiological marker of autonomic and brainstem recovery, highlighting the need for further longitudinal investigation.

## Data Availability

Due to the clinical nature of the data, they are subject to privacy/ethical restrictions, and are available to researchers only upon request by contacting the corresponding author.

## Acknowledgments

We would like to express our sincere gratitude to the caregivers at HCL (Hospices Civils de Lyon) for their support. We also thank the computer scientists, Hervé Hugueney and Thibaut Woog, at CRNL (Centre de Recherche en Neurosciences de Lyon) for developing the valuable computing environment that enabled us to process large data sets.

## Data availability

The datasets reported herein are available from the corresponding author on a reasonable request.

## Code availability

The code used in this study includes contributions from multiple sources:

- **physio Toolbox**: Available at https://github.com/samuelgarcia/physio
- **pycns Toolbox**: Available at https://github.com/samuelgarcia/pycns
- **Project-Specific Code**: Developed for this study and available at https://github.com/ValentinGhibaudo/RespHRV_NeuroICU

## Fundings

Contrat CCA Inserm-Bettencourt, Fondation des Gueules Cassées, Hospices Civils de Lyon

## Authors statement

Initials: VG, GP, SG, HA, NB, CM, BB

- Conception and Design: VG, BB
- Data Collection: VG, GP, NB, BB
- Data Analysis and Interpretation: VG, SG
- Methodology Development: VG, SG, BB
- Manuscript Drafting and Writing: VG, HA, BB
- Supervision and Project Oversight: BB
- Funding Acquisition: BB, VG, NB
- Visualization: VG, SG
- Critical Review and Editing: HA, NB, BB, CM
- Approval of the Final Version: All
- Software Development: VG, SG

## Abbreviations

ICU: Intensive Care Unit
GCS: Glasgow Coma Scale
mRS: Modified Rankin Scale
SAH: Subarachnoid Hemorrhage
TBI: Traumatic Brain Injury
ABP: Arterial Blood Pressure
ECG: Electrocardiogram
HRV: Heart Rate Variability
RespHRV: Respiratory Heart Rate Variability
rCPG: Central Pattern Generators of Respiration
HR: Heart rate

## Notes

### Competing Interest Statement

The authors have declared no competing interest.

### Funding Statement

Contrat CCA Inserm-Bettencourt, Fondation des Gueules Cassees, Hospices Civils de Lyon

### Author Declarations

The MultiICU study was approved by the Comité scientifique et éthique des Hospices Civils de Lyon (Ethical Committee IRB 0013204, No. 713, 06/07/2023) and registered with the French National Commission on Informatics and Liberty (CNIL No. 25_5713, France).

